# Implementation and Evaluation of a Postoperative Virtual Hospital Pathway for Elective Colorectal Surgery: A Propensity-Matched Analysis

**DOI:** 10.64898/2026.05.20.26353655

**Authors:** Harry Whelan, Bibechan Thapa, Samuel Massias, Lillian Reza, Nicholas Johnson, James Kinross, Najib Daulatzai, Vanash Patel

**Author notes:** Correspondence to: Mr Vanash M Patel, Consultant Colorectal Surgeon, Department of Colorectal Surgery, West Hertfordshire Teaching Hospitals NHS Trust, Watford General Hospital, Vicarage Road, WD18 0HB. No funding/support. No financial disclosures. Author Contributions: **Harry Whelan** contributed to study conception and design, acquisition of data, data analysis and interpretation, drafting of the manuscript, and critical revision for important intellectual content. He approved the final version for publication. **Bibechan Thapa** contributed to acquisition of data, data analysis and interpretation, drafting of the manuscript, and critical revision for important intellectual content. He approved the final version for publication. **Samuel Massias** contributed to acquisition of data, data analysis and interpretation, drafting of the manuscript, and critical revision for important intellectual content. He approved the final version for publication. **Lillian Reza** contributed to study conception and design, acquisition of data, interpretation of results, drafting of the manuscript, and critical revision for important intellectual content. She approved the final version for publication. **Nicholas Johnson** contributed to study design, statistical methodology, data analysis and interpretation, and critical revision of the manuscript for important intellectual content. He approved the final version for publication. **James Kinross** contributed to study conception and design, interpretation of data, and critical revision of the manuscript for important intellectual content. He approved the final version for publication. **Najib Daulatzai** contributed to study conception and design, acquisition and interpretation of data, and critical revision of the manuscript for important intellectual content. He approved the final version for publication. **Vanash Patel** conceived and designed the study, led data acquisition, analysis and interpretation, drafted the manuscript, provided critical revision for important intellectual content, and supervised the project. He approved the final version for publication and is the guarantor of the work.

## Abstract

**Background:** Virtual Hospital (VH) pathways enable early discharge and are promoted across the NHS. However, evidence supporting their safety, effectiveness, equity, and patient acceptability in elective colorectal surgery remains limited.

**Objective:** To evaluate implementation of a VH pathway following elective colorectal surgery and its impact on clinical outcomes, patient-centred recovery, and equality, diversity and inclusion (EDI).

**Design:** Retrospective service evaluation with a historical comparator. A 1:1 propensity-matched analysis was performed in colorectal cancer patients using age, sex, body mass index, ASA grade, Charlson Comorbidity Index, procedure type, tumour site, and anastomosis.

**Setting:** West Hertfordshire Teaching Hospitals NHS Trust.

**Patients:** Patients undergoing elective colorectal surgery between November 2023 and March 2025 managed via a VH pathway, compared with a non-VH cohort.

**Main outcome measures:** Primary outcomes were inpatient length of stay (IPLOS), VH length of stay (VHLOS), and days alive and at home within 30 days (DAH30). Secondary outcomes were patient-reported experience and EDI characteristics.

**Results:** Eighty-one patients were managed via VH. Median [Q1–Q3] IPLOS was 2 [1–2] days and VHLOS was 2 [2–3] days, with no deaths. Forty-two colorectal cancer patients were propensity matched 1:1 to a pre-VH cohort. IPLOS was shorter in the VH cohort (2 [1–2] vs 4 [3–5] days; p<0.001), and DAH30 was higher (28 [28–29] vs 25.5 [24–27] days;p<0.0001). Patient experience was positive, with mean satisfaction >8.5/10 and over 90% preferring VH-supported recovery. Sex and ethnicity distributions were similar, although VH patients were younger (p=0.028).

**Limitations:** This single-centre retrospective evaluation had a modest sample size and non-randomised design. VH patients were carefully selected, limiting generalisability.

**Conclusions:** A VH pathway following elective colorectal surgery is feasible, safe, and acceptable. Compared with a pre-VH cohort, VH care reduced inpatient stay and improved patient-centred recovery without compromising safety.

## INTRODUCTION

The National Health Service (NHS) Delivery Plan for Recovering Urgent and Emergency Care Services in 2023 established Virtual Hospital (VH) (also known as Virtual Wards or Hospital at Home) as an efficient and safe alternative to inpatient care to meet the demands of rising patient volumes, resource constraints, and the need for equitable access^1^. By 2023, the NHS had 15,000 virtual beds and aims to expand capacity to 25,000 using remote monitoring and digital technology, with oversight from a multidisciplinary hospital-at-home care team. The key aims are to manage health in the comfort and convenience of home and to avoid hospital readmissions. Despite dedicated funding and guidance for implementation of VH, virtual bed occupancy was approximately 76% in 2025^2^. Although VH pathways are resource-intensive and require careful patient selection, emerging real-world NHS evidence has demonstrated reductions in inpatient bed utilisation, readmissions, and overall healthcare costs following implementation of hospital-at-home services^3^.

To date, few surgical studies have reported data supporting the national uptake of VH to streamline the early and safe discharge of elective surgical patients. Globally, the only randomised controlled trial (RCT) data for VH in surgery is being generated by the Canadian Post Discharge After Surgery Virtual Care with Remote Automated Monitoring Technology studies^4–6^. PVC-RAM-1 reported favourable outcomes of remote monitoring in patients following non-elective surgery, including a reduction in hospital readmissions^5^. Neither PVC-RAM-2 (semi-urgent (e.g., oncology), urgent or emergency surgery) nor PVC-RAM-3 (adults undergoing elective non-cardiac surgery) has yet reported their data^4,6^.

Elective colorectal surgery represents a prime opportunity for innovation in VH pathways. Enhanced Recovery After Surgery (ERAS) perioperative protocols have become standard and have improved postoperative recovery and reduced length of stay nationally^7^. VH represents a natural extension of this strategy, with the potential to strengthen Integrated Care Systems, support the NHS’s population health agenda and improve access to high-quality cancer surgery^8,9^. Few VH studies have reported patient satisfaction as an outcome, and the certainty of the evidence is considered low^10^.

Implementing a VH pathway in colorectal surgery introduces distinct challenges that extend beyond technological considerations. Surgeons are characteristically risk-averse, particularly due to the potential for critical complications such as anastomotic leak^11^. The lack of definitive evidence supporting the safety and cost-effectiveness of early discharge with virtual care within this patient population has hindered the implementation of VH in colorectal surgery.

We report the implementation of a VH pathway designed to facilitate early discharge in carefully selected patients undergoing elective colorectal surgery. This pilot analysis evaluated its safety, feasibility, and impact on patient experience.

## METHODS

### Ethics approval

This study was conducted as a service evaluation of an established postoperative virtual hospital pathway within West Hertfordshire Teaching Hospitals NHS Trust (WHTH). In accordance with UK Health Research Authority guidance, the project was not classified as research because its primary aim was to evaluate and refine an existing clinical service rather than generate generalisable knowledge or alter patient care pathways. Consequently, formal HRA or NHS Research Ethics Committee approval was not required.

The project was reviewed and approved locally by the Information Governance team and the Clinical Lead for Surgery at WHTH prior to commencement. Patients included in the pathway had provided consent for clinical care and participation in the virtual hospital pathway as part of routine service delivery.

### Patient recruitment

All patients included in the study underwent surgery at Watford General Hospital, part of WHTH, which performs approximately 250 major elective colorectal resections annually for benign and malignant disease. Prior to 2023, the mean length of stay (LOS) was 6.5 days^12^. In November 2023, a VH pathway was launched to support early discharge with remote monitoring in carefully selected patients. Based on pilot modelling, approximately 15% of patients were expected to be eligible for discharge within 24–72 hours postoperatively.

Eligibility was assessed in an outpatient clinic or at the colorectal multidisciplinary team (MDT) meeting. Patients were counselled by the consultant colorectal surgeon, reviewed in the preoperative assessment clinic, and received additional guidance from the ERAS nurse specialist. An information booklet and in-person training were provided to patients and caregivers on device use and postoperative care. Eligibility criteria are summarised in **Table 1**.

**Table 1:** Eligibility criteria for VH.

| **Pre-operative eligibility criteria** | **Post-operative eligibility criteria** |
| --- | --- |
| ASA I–II and $\leq 75$ years | No new stoma formed |
| Minimally invasive colorectal resections (laparoscopic/robotic) and stoma reversals<br>Operation not expected to require a new stoma | Observations stable with no evidence of deterioration (e.g. NEWS2 $\leq 4$ and no score of 3 in a single parameter) |
| Reliable support to return to hospital urgently if needed (family/friends or community/VH support) | No increasing oxygen requirement and no significant organ dysfunction |
| No complex social, safeguarding, cognitive, or mobility issues preventing safe home monitoring | Post-operative haemoglobin demonstrating no significant blood loss and normal renal function |
| Able to understand and use (or be supported to use) the required monitoring technology/equipment | Pain controlled with oral analgesia |
|  | Tolerating soft diet without vomiting |
|  | Not requiring intravenous fluids or intravenous medications |
|  | Able to self-administer prophylactic enoxaparin, or family/carer trained to administer |

Following surgery, patients were jointly assessed by the consultant surgeon, consultant anaesthetist, and ERAS/ward nurse. Those meeting VH admission criteria were discharged with a personalised treatment plan and enrolled in remote monitoring.

The VH pathway was implemented as part of routine clinical care and service development within the institution rather than as an interventional research study. VH therefore represented a standard clinical pathway for selected patients considered suitable for early supported discharge following colorectal surgery. Participation in the VH pathway was voluntary, and eligible patients were offered the option of conventional inpatient postoperative care if they did not wish to participate in remote monitoring or early discharge. Patients received verbal and written information regarding the pathway during the preoperative assessment process, including the rationale for remote monitoring, escalation procedures, and follow-up arrangements.

### Virtual hospital framework

For this study, a VH was defined as “an acute clinical service with staff, equipment, technologies, medication and skills usually provided in hospitals, delivered to selected patients in their usual place of residence, including care homes”.

Prior to discharge, patients were onboarded by the nursing team and given written guidance on managing common postoperative issues. Monitoring included a Masimo™ pulse oximeter (oxygen saturation, pulse) and a blood pressure device, with data transmitted via Bluetooth® to a tablet four times daily and integrated into the electronic patient record. Patients also received daily telephone follow-up from a VH ‘hub’ nurse and a telehealth consultation with a surgical team member to review recovery, track milestones, and confirm readiness for discharge from VH (**see Supplementary Material S1**).

Escalation pathways included in-person review in the surgical acute care clinic (08:00– 17:00), the surgical assessment unit (17:00–08:00), or direct hospital admission, if required. Patient flow through the VH pathway is outlined in **Figure 1**.

**Figure 1:**
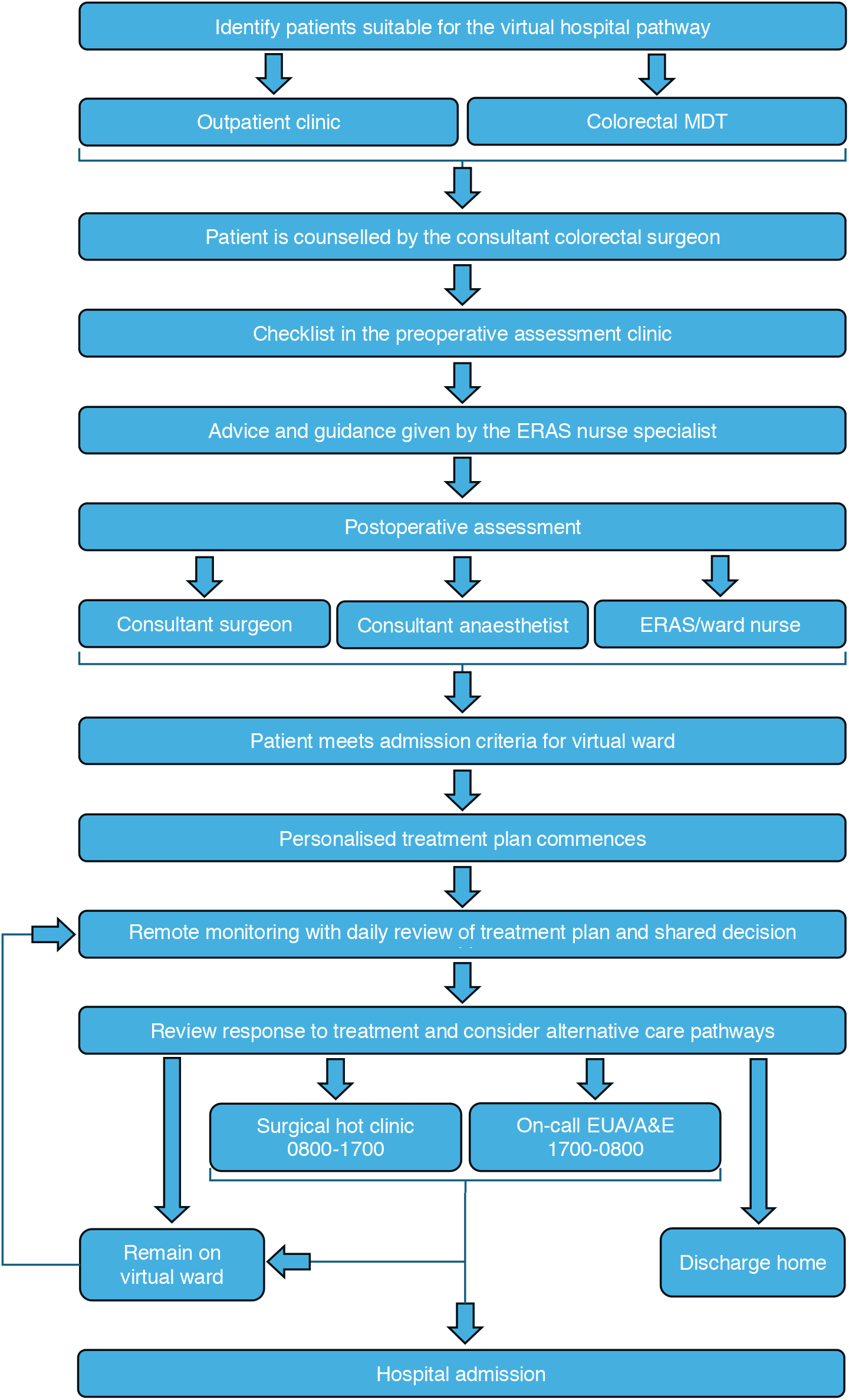
Flowchart showing the VH pathway

The VH service was staffed by a multidisciplinary team comprising two colorectal consultants, one ERAS specialist nurse, four clinical nurse specialists, ten ward nurses, four administrative staff, and one pharmacist. Staffing for the pilot pathway was delivered using existing personnel from the colorectal surgical and established virtual hospital services, without dedicated additional staffing appointments specifically for the study.

### Primary outcomes

A retrospective dataset was extracted from Cerner Millennium™, the electronic health record system at WHTH. All patients enrolled on the VH pathway between November 2023 and March 2025 were identified. For the pre-VH comparator group, patients undergoing elective minimally invasive colorectal cancer resection between November 2021 and October 2023 were included.

Clinical data were obtained from electronic patient records and included patient demographics, diagnosis and staging, preoperative comorbidities, operative details (procedure type and duration), estimated blood loss, perioperative or postoperative complications, length of hospital stay and readmission stay. Data extraction was performed by HW and SM, with all data independently validated for accuracy by VP. Days Alive and at Home within 30 days (DAH30) was calculated as a composite measure reflecting postoperative recovery, incorporating length of stay, readmissions and mortality^13^.

### Secondary outcomes

#### Patient experience

Patient-reported outcome measures were collected using structured questionnaires administered to all patients at enrolment into the VH pathway and at one week following discharge. Due to service-delivery constraints during implementation, patient experience data were collected using two sequential survey instruments.

During the first six months, an in-house questionnaire was used to assess overall satisfaction, measured on an 11-point Likert scale, and care preference, captured as a binary response comparing the VH pathway with continued inpatient care. Additional domains included perceived usefulness of the remote monitoring devices, clarity of the Masimo SafetyNet™ application interface (categorical ratings), and perceived helpfulness of the central monitoring hub. The questionnaire demonstrated face validity and clinical relevance; however, formal psychometric validation was not undertaken due to the pilot nature of the study. Patients were also invited to provide free-text feedback on the usability and acceptability of the VH service. From month six onward, the IQVIA™ patient experience survey—a validated instrument developed by a global provider of advanced analytics, technology solutions, and clinical research services—was adopted to reduce the administrative burden on the VH team (**see Supplementary Material S2**). This tool generates mean scores for each question, which are reported accordingly. While both instruments assessed comparable domains of patient experience, minor methodological differences existed; therefore, results from each survey were analysed and reported separately.

#### Equality, diversity and inclusion measures

Equality, diversity and inclusion were assessed by systematically screening all inpatient admissions on the colorectal waiting list between November 2023 and March 2025. Case identification was undertaken by a VH data analyst in collaboration with the study team. A predefined set of primary OPCS-4 (Office of Population Censuses and Surveys Classification of Surgical Operations and Procedures, version 4) procedure codes representing colorectal resections and stoma reversals (**see Supplementary Material S3**) was applied to extract eligible admissions from the Cerner data warehouse.

Each identified admission was cross-referenced with the VH recruitment database to determine inclusion in the VH pathway, with unmatched patients classified as non-VH comparators. Equality, diversity and inclusion measures, including age, sex and ethnicity, were extracted for all included patients. These measures were used to assess representativeness across key protected characteristics defined under the Equality Act 2010.

### Statistical analysis

Patients undergoing bowel resection and stoma reversal demonstrated distinct perioperative and recovery trajectories, particularly regarding bowel function recovery, complication profiles, and duration of VH monitoring, and were therefore analysed descriptively rather than directly compared. Using data from patients who had colorectal cancer, post-hoc propensity score matching was undertaken to create two comparative cohorts—with and without a VH pathway. Of the 81 VH patients, a subgroup of 43 treated for colorectal adenocarcinoma was matched to a retrospective cohort of 254 patients who underwent surgery before VH implementation. An optimal fixed ratio (1:1) approach was used with scores derived using variables age, sex, body mass index (BMI), American Society of Anaesthesiologists (ASA) grade, Charlson Comorbidity Index (CCI), operation type, tumour site (colon or rectal) and anastomosis. Following successful matching, the cohorts were evaluated across demographic and cancer aetiology parameters to confirm balance between groups. LOS and DAH30 were assessed using a Poisson regression model with a log-link function, while rates of complications, anastomotic leak, unplanned intensive care unit admission, and Clavien–Dindo ≥ II complications were analysed using logistic regression. Data were processed and analysed using SAS (Version 9.4).

## RESULTS

### Primary outcomes

A total of 81 patients were managed via the VH. **Table 2** summarises the preoperative, perioperative, and postoperative characteristics of this cohort. Within this group, patients were managed through either a bowel resection pathway (n=54) or a stoma reversal pathway (n=27).

**Table 2:** Preoperative, perioperative, and postoperative characteristics following bowel resection or stoma reversal in the VH cohort. Continuous variables are presented as median [Q1-Q3], and categorical variables as n (%).

| **BOWEL RESECTION** | 54 (67%) | **STOMA REVERSAL** | 27 (33%) |
| --- | --- | --- | --- |
| **PREOPERATIVE** |  | **PREOPERATIVE** |  |
| **Baseline characteristics** |  | **Baseline characteristics** |  |
| Male | 27 (50%) | Male | 20 (74%) |
| BMI, kg/m2 | 27 [25-29] | BMI, kg/m2 | 28 [25-31] |
| ASA | 2 [2-2] | ASA | 2 [2-2] |
| **Diagnosis** |  |  |  |
| Cancer | 48 (89%) |  |  |
| Inflammatory bowel disease | 2 (4%) |  |  |
| Diverticular disease | 2 (4%) |  |  |
| Benign polyp | 2 (4%) |  |  |
| **PERIOPERATIVE** |  | **PERIOPERATIVE** |  |
| **Operation type** |  | **Operation type** |  |
| Laparoscopic small bowel resection | 4 (7%) | Closure of colostomy | 3 (11%) |
| Robotic anterior resection | 17 (31%) | Closure of Ileostomy | 24 (89%) |
| Robotic right hemicolectomy | 25 (46%) |  |  |
| Robotic abdominoperineal resection | 1 (2%) |  |  |
| Laparoscopic right hemicolectomy | 1 (2%) |  |  |
| Robotic sigmoid colectomy | 2 (4%) |  |  |
| Open reversal of hartmann's | 1 (2%) |  |  |
| Robotic posterior pelvic exenteration | 1 (2%) |  |  |
| Robotic left hemicolectomy | 2 (4%) |  |  |
| **POSTOPERATIVE** |  | **POSTOPERATIVE** |  |
| **Indwelling devices** |  | **Indwelling devices** |  |
| Abdominal drain | 18 (33%) | Abdominal drain | 0 (0%) |
| Nasogastric tube | 0 (0%) | Nasogastric tube | 0 (0%) |
| Urinary catheter | 50 (93%) | Urinary catheter | 0 (0%) |
| **Analgesia** |  | **Analgesia** |  |
| Epidural | 4 (7%) | Epidural | 0 (0%) |
| Patient controlled analgesia | 38 (70%) | Patient controlled analgesia | 2 (7%) |
| **Oncological outcomes** |  |  |  |
| R0 resection | 48 (100%) |  |  |
| Lymph node yield | 23 [IQR 18-31] |  |  |

Median [Q1–Q3] inpatient length of stay (IPLOS) was 2 days [1–2] and the VH length of stay (VHLOS) was 2 days [2–3]. Thirty-day readmission rate was 7.4% (6 patients). Bowel resections had a median IPLOS of 2 [1–2] days and VHLOS of 2 [2–3] days, with a 30-day readmission rate of 3.7% (2 patients). For stoma reversals, median IPLOS was 1 [1–2] days and VHLOS was 3 [2–4] days, with a 30-day readmission rate of 14.8% (4 patients).

The median time to readmission from VH discharge was 4.5 [0.25-11.75] days. Among these six patients readmitted from the VH cohort, two were diagnosed with ileus, one experienced mild rectal bleeding, one tested positive for norovirus, and two presented with pyrexia that resolved with short courses of antibiotics after a two-day hospital stay. Our VH monitoring program detected two complications. The other four complications that necessitated readmission occurred after discharge from VH. No deaths occurred in the VH cohort.

### Propensity-matched outcomes

Of the 43 eligible VH patients (colorectal adenocarcinoma), 42 were successfully matched 1:1 to patients within the non-VH dataset. All binary variables within the model achieved negligible standardised mean differences. Non-binary model variables; age, BMI, and operation type were assessed manually but also indicated balance, demonstrating that comparable cohorts were attained. Subsequent review indicated that baseline demographic characteristics were well matched between cohorts. However, a modest imbalance was observed in the proportion of patients from ethnic minority backgrounds (13 [31%] in the VH cohort vs 6 [14%] in the non-VH cohort). Cancer aetiology was comparable across groups. **Table 3** describes the baseline characteristics, operation type, pathological staging and ethnicity in the propensity-matched VH cohort and pre-VH comparator cohort.

**Table 3:** Baseline characteristics, operation type, pathological staging and ethnicity in the propensity-matched VH cohort and pre-VH comparator cohort. Continuous variables are presented as median [Q1-Q3], and categorical variables as n (%).

| Variable | VH (n=42) | Pre-VH (n=42) |
| --- | --- | --- |
| **Baseline characteristics** |  |  |
| Age, years | 68 [58–73] | 70 [60–76] |
| Male | 21 (50.0%) | 19 (45.2%) |
| BMI, kg/m<sup>2</sup> | 26.0 [24.0–28.0] | 26.7 [23.1–28.3] |
| ASA | 2 [2–2] | 2 [2–2] |
| CCI score | 5 [4–6] | 5 [4–6] |
| **Operation type** |  |  |
| Right-sided colectomy | 22 (52.4%) | 23 (54.8%) |
| Left-sided colectomy | 2 (4.8%) | 2 (4.8%) |
| Rectal resection | 18 (42.9%) | 17 (40.5%) |
| Anastomosis | 40 (95.2) | 39 (92.9%) |
| **Pathological staging** |  |  |
| T stage |  |  |
| T0 | 2 (4.8%) | 1 (2.4%) |
| T1 | 8 (19.0%) | 1 (2.4%) |
| T2 | 8 (19.0%) | 11 (26.2%) |
| T3 | 20 (47.6%) | 24 (57.1%) |
| T4 | 4 (9.5%) | 5 (11.9%) |
| N stage |  |  |
| N0 | 25 (59.5%) | 31 (73.8%) |
| N1 | 10 (23.8%) | 6 (14.3%) |
| N2 | 7 (16.7%) | 5 (11.9%) |
| M stage |  |  |
| M0 | 41 (97.6%) | 39 (92.9%) |
| M1 | 1 (2.4%) | 3 (7.1%) |
| **Ethnicity** |  |  |
| Ethnic minority | 13 (31.0%) | 6 (14.3%) |

**Table 4:** Comparison of ethnic distribution between VH and Non-VH cohorts.

| **Ethnicity** | **VH** | **Non-VH** |
| --- | --- | --- |
| White – British | 56 | 199 |
| White – Any Other White Background | 7 | 13 |
| White – Irish | 3 | 6 |
| Asian – Indian | 1 | 5 |
| Asian – Bangladeshi | 0 | 3 |
| Asian – Other Asian Background | 1 | 3 |
| Pakistani | 0 | 1 |
| Black – Caribbean | 1 | 3 |
| Black – Other Black Background | 0 | 3 |
| Mixed – Any Other Mixed Background | 0 | 2 |
| Chinese | 0 | 1 |
| Other – Any Other Ethnic Group | 0 | 3 |
| Not Stated | 12 | 11 |

In the matched analysis, median [Q1–Q3] IPLOS was significantly shorter in the VH cohort at 2 [1–2] days compared with 4 [3–5] days in the non-VH cohort (p < 0.001). The median duration of VH monitoring was 2 [2–3] days, resulting in a combined total length of stay of 4 [3–6] days for VH patients. Days alive and at home at 30 days (DAH30) was significantly higher in the VH cohort at 28 [28–29] days compared with 25.5 [24–27] days in the non-VH cohort (p < 0.0001).

Postoperative outcomes were similar between cohorts. Rates of any complication were 9 (21%) in the VH group versus 11 (26%) in the non-VH group; anastomotic leaks occurred in 0 (0%) versus 1 (2%) patients, and unplanned ICU admissions in 0 (0%) versus 1 (2%), respectively. Clavien–Dindo grade ≥II complications were observed in 8 (19%) of VH patients and 10 (24%) of non-VH patients. Model-adjusted logistic regression demonstrated no statistically significant between-group differences for any complication, anastomotic leak, unplanned ICU admission, or Clavien–Dindo ≥II complications (p = 0.61, 0.12, 0.31, and 0.59, respectively).

### Secondary outcomes

#### Patient experience

Thirty-three patients (41%) in the VH cohort responded to the patient experience surveys, which included both the original questionnaire and the IQVIA-administered survey.

A total of 22 patients (27%) completed the original patient experience questionnaire. Feedback was positive, with a mean satisfaction score of 8.5 out of 10. Notably, 95% (21/22) of respondents felt that the VH pathway was preferable to continued inpatient care, while 100% (22/22) reported feeling safe during recovery at home. Engagement with the remote monitoring hub was also well received: 95% (21/22) found the hub team helpful, and the same proportion felt their concerns or questions were adequately addressed. Ease of use was a core design priority for the VH programme. Encouragingly, 91% (20/22) found the clinical monitoring devices easy to use, and 77% (17/22) rated the Masimo SafetyNet application interface as easy to navigate.

An additional 11 patients (14%) responded to the IQVIA experience survey. In this cohort, the mean satisfaction score was 8.81 out of 10. Most patients (86.4%) reported being glad to recover in a VH bed rather than remain in hospital. The majority (79.5%) felt safe throughout their stay on the VH pathway. Furthermore, 81.8% of patients stated that the remote monitoring hub team understood their needs, and the same proportion felt well informed about their health during their virtual stay. In terms of usability, 85% of patients reported that it was easy to record their observations using the clinical monitoring devices, and 78.6% of those who used the Masimo application found it easy to use.

#### Equality, diversity and inclusion measures

A total of 334 patients were included in the Cerner dataset, comprising 81 managed via the VH and 253 managed conventionally (Non-VH). Patients in the VH cohort were significantly younger, with a median [IQR] age of 62 years [52–71] compared with 66 years [55–76] in the Non-VH group (p=0.028). The sex distribution was comparable between cohorts (VH: 59% male vs Non-VH: 60% male, p=1.000). Ethnic composition is reported in the **Supplementary Material S4**.

## DISCUSSION

### Principal findings

This pilot analysis suggests that implementation of a VH pathway following elective colorectal surgery is feasible and appears safe, with meaningful improvements in both healthcare utilisation and patient-centred recovery^14^. In addition to substantial reductions in inpatient length of stay, VH care was associated with a significant increase in DAH30 compared with a pre-VH cohort. DAH30 is an increasingly recognised composite outcome that captures not only hospital stay but also readmissions and early mortality and therefore provides a more holistic measure of postoperative recovery^15^. The observed improvement in DAH30 supports the premise that early discharge supported by remote monitoring can permit recovery at home without compromising safety. These findings are consistent with prior VH and remote monitoring studies, which have demonstrated improved efficiency and recovery across a range of surgical and medical settings, and further reinforce the role of digitally enabled pathways as an extension of enhanced recovery programmes^16,17^.

### Strengths and weaknesses of the study

Key strengths of this study include its real-world NHS implementation, use of DAH30 as a patient-centred composite outcome, and incorporation of patient experience and EDI analyses alongside traditional clinical metrics. Propensity matching within the colorectal cancer subgroup strengthened comparability with historical controls.

However, several limitations must be acknowledged. As a pilot service evaluation, the sample size was small and the study lacked randomisation or a concurrent control group, limiting causal inference. Although propensity matching improved comparability between cohorts, the use of historical controls introduces potential temporal confounding, highlighting the need for prospective randomised studies comparing VH-supported care with standard postoperative pathways. The VH pathway was introduced alongside increased adoption of robotic colorectal surgery, which itself may improve postoperative recovery and reduce length of stay. Consequently, operative approach represents a potential confounder, and the observed benefits may reflect the combined effects of minimally invasive surgery and VH-supported care rather than VH alone. The cohort largely comprised low-risk patients, and those with stomas were excluded due to community support constraints, restricting generalisability. Patient experience data were derived from a small, self-selected subgroup with a low response rate, introducing potential response bias and limiting the interpretability of the patient-reported outcomes, particularly as two different survey instruments were used during the implementation period. These findings should therefore be interpreted primarily as evidence of feasibility rather than definitive effectiveness.

### Strengths and weaknesses in relation to other studies

The observed reductions in length of stay and acceptable readmission rates are consistent with the Canadian PVC-RAM-1 evaluations of VH and virtual ward models^3^. Compared with this study, the present analysis was smaller and relied on historical comparators rather than contemporaneous controls, limiting direct comparison. However, this study adds to the literature by demonstrating feasibility within a UK NHS colorectal service and by incorporating DAH30 and patient-reported experience, outcomes that are often underreported in earlier VH evaluations.

### Meaning of this study

The benefits observed with VH care are likely driven by a combination of factors, including earlier mobilisation and improved patient engagement, rather than early discharge alone. Although some patients undergoing minimally invasive colorectal surgery may be suitable for discharge with standard outpatient follow-up alone, postoperative length of stay in the UK remains variable and is often influenced by caution regarding recovery of bowel function and early detection of complications. The VH pathway was therefore initially implemented as a proof-of-concept service in carefully selected lower-risk patients before expansion to more complex cohorts. VH pathways may function most effectively as an extension of ERAS programmes, supporting recovery at home while maintaining clinical oversight.

From a systems perspective, reduced inpatient length of stay has important implications for bed capacity and patient flow. Importantly, patients valued shorter hospitalisation, emphasising that VH pathways align clinical efficiency with patient priorities. However, VH implementation must be accompanied by clear escalation pathways for face-to-face review, recognising that not all postoperative complications can be detected remotely.

### Unanswered questions and future research

Several questions remain. Larger, multicentre controlled studies are required to confirm clinical effectiveness, cost-effectiveness, and scalability. Comparative studies evaluating VH-supported care against early discharge pathways without VH monitoring would help define the specific utility and added value of remote postoperative surveillance. Future evaluation should also focus on identifying which patient subgroups derive the greatest benefit and how VH pathways can be adapted for older, higher-risk, or digitally excluded populations to avoid widening health inequities. Integration of social care and community services within VH models represents a promising but underexplored area, particularly for patients with multimorbidity or limited support at home. Addressing these gaps will be essential to ensure that VH pathways are both effective and equitable as they expand.

## Data Availability

All data produced in the present study are available upon reasonable request to the authors.

## ACKNOWLEDGEMENTS

The authors thank Laura Payne and Deirdre McCarthy for their essential contributions to the implementation and day-to-day delivery of the Virtual Hospital pathway at West Hertfordshire Teaching Hospitals NHS Trust.

## REFERENCES

1. Getting It Right First Time. New summary guide supports NHS ambition to increase the use of virtual wards [Internet]. 2023 [cited 2025 Oct 22]. Available from: https://gettingitrightfirsttime.co.uk/new-summary-guide-supports-nhs-ambition-to-increase-the-use-of-virtual-wards/

2. Cathers S, Lally C. Virtual wards and hospital at home [Internet]. 2025 [cited 2025 Oct 22]. Available from: https://researchbriefings.files.parliament.uk/documents/POST-PN-0744/POST-PN-0744.pdf

3. Shaw M, Almogheer B, Auger D, Barlow A, Bhaskaran B, Buxton M, et al. Real-world outcomes from 2,905 episodes of hospital at home care: a propensity-matched cohort study. Front Digit Health. 2026 Apr 8;8:1716319. doi:10.3389/fdgth.2026.1716319

4. McGillion MH, Parlow J, Borges FK, Marcucci M, Jacka M, Adili A, et al. Post Discharge after Surgery Virtual Care with Remote Automated Monitoring Technology (PVC-RAM): protocol for a randomized controlled trial. cmajo. 2021 Jan;9(1):E142–8. doi:10.9778/cmajo.20200176

5. McGillion MH, Parlow J, Borges FK, Marcucci M, Jacka M, Adili A, et al. Postdischarge after surgery Virtual Care with Remote Automated Monitoring-1 (PVC-RAM-1) technology versus standard care: randomised controlled trial. BMJ. 2021 Sep 30;n2209. doi:10.1136/bmj.n2209

6. Ofori S, McGillion MH, Borges FK, Ouellette C, Patel A, Conen D, et al. Impact of Virtual Care With Remote Automated Monitoring on the Rate of Acute Hospital Care Post Discharge and Index Length of Hospital Stay: Protocol for the Post Discharge After Surgery Virtual Care With Remote Automated Monitoring Technology 3 (PVC-RAM-3) Trial. JMIR Res Protoc. 2025 Jun 2;14:e72672. doi:10.2196/72672

7. Telem DA, Sur M, Tabrizian P, Chao TE, Nguyen SQ, Chin EH, et al. Diagnosis of gastrointestinal anastomotic dehiscence after hospital discharge: Impact on patient management and outcome. Surgery. 2010 Jan;147(1):127–33. doi:10.1016/j.surg.2009.06.034

8. Braun M, Fearnhead N, Walker K, Cook S, van der Meulen J, Kuryba A, et al. National Bowel Cancer Audit State of the Nation Report [Internet]. 2025 [cited 2025 Oct 22]. Available from: https://www.natcan.org.uk/wp-content/uploads/2025/10/NBOCA-State-of-the-Nation-Report-2025.pdf

9. Feng Q, Yuan W, Li T, Tang B, Jia B, Zhou Y, et al. Robotic versus laparoscopic surgery for middle and low rectal cancer (REAL): short-term outcomes of a multicentre randomised controlled trial. The Lancet Gastroenterology & Hepatology. 2022 Nov;7(11):991–1004. doi:10.1016/S2468-1253(22)00248-5

10. McGrath J, Briggs T, Vig S. Implementation of robotic-assisted surgery (RAS) in England [Internet]. 2025 [cited 2025 Oct 22]. Available from: https://gettingitrightfirsttime.co.uk/wp-content/uploads/2025/07/FINAL_NHS-England-and-GIRFT-implementation-of-robotically-assisted-surgery-in-England_17-07-2025.pdf

11. Millions to benefit from NHS robot drive [Internet]. 2025. Available from: https://www.england.nhs.uk/2025/06/millions-to-benefit-from-nhs-robot-drive/

12. Massias S, Vadhwana B, Arjomandi Rad A, Hollingshead J, Patel V. Feasibility, clinical outcomes, and learning curves of robotic-assisted colorectal cancer surgery in a high-volume district general hospital: a cohort study. Annals of Medicine & Surgery. 2024 Oct;86(10):5744–9. doi:10.1097/MS9.0000000000002545

13. Myles PS, Shulman MA, Heritier S, Wallace S, McIlroy DR, McCluskey S, et al. Validation of days at home as an outcome measure after surgery: a prospective cohort study in Australia. BMJ Open. 2017 Aug 18;7(8):e015828. doi:10.1136/bmjopen-2017-015828 PubMed PMID: 28821518; PubMed Central PMCID: PMC5629653.

14. Virtual wards enabled by technology: Guidance on selecting and procuring a technology platform [Internet]. [cited 2025 Aug 12]. Available from: https://www.england.nhs.uk/long-read/virtual-wards-enabled-by-technology-guidance-on-selecting-and-procuring-a-technology-platform/

15. Groff AC, Colla CH, Lee TH. Days Spent at Home - A Patient-Centered Goal and Outcome. N Engl J Med. 2016 Oct 27;375(17):1610–2. doi:10.1056/NEJMp1607206 PubMed PMID: 27783911; PubMed Central PMCID: PMC5996758.

16. Steel RP, Field JD, Dieleman JM. The value of Days Alive and Out of Hospital 30 Days After Surgery as an outcome measure in a Major Cardiac Surgical Centre in Australia. JTCVS Open. 2025 Dec;101556. doi:10.1016/j.xjon.2025.101556

17. Ariti CA, Cleland JGF, Pocock SJ, Pfeffer MA, Swedberg K, Granger CB, et al. Days alive and out of hospital and the patient journey in patients with heart failure: Insights from the Candesartan in Heart failure: Assessment of Reduction in Mortality and morbidity (CHARM) program. American Heart Journal. 2011 Nov;162(5):900–6. doi:10.1016/j.ahj.2011.08.003

