## Supplementary Material for "Implementation and Evaluation of a Postoperative Virtual Hospital Pathway for Elective Colorectal Surgery: A Propensity-Matched Analysis"

Harry Whelan[1] MBBS, Bibechan Thapa[1] MBBS MRCS, Samuel Massias[1] MBBS MRCS, Lillian Reza[1] MBBS MSc FRCS, Laura Payne[1], Deirdre McCarthy[1], Nicholas Johnson[3], James Kinross[2] MBBS PhD DIC FRCS, Najib Daulatzai[1] MBBS MD FRCS, Vanash Patel[1,2] MBBS MSc PhD DIC FRCS

Affiliations:

1. Department of Surgery, West Hertfordshire Teaching Hospitals NHS Trust, Watford General Hospital, Vicarage Road, WD18 0HB, UK.
2. Department of Surgery and Cancer, Imperial College London, 10th Floor QEQM Building, St Mary's Hospital, London W2 1NY, UK.
3. Imperial Clinical Trials Unit (ICTU), Imperial College London, 1st Floor, Stadium House, 68 Wood Lane, London W12 7RH, UK.

Correspondence to:
Mr Vanash M Patel, Consultant Colorectal Surgeon, Department of Colorectal Surgery, West Hertfordshire Teaching Hospitals NHS Trust, Watford General Hospital, Vicarage Road, WD18 0HB


#### Supplementary Methods

## S1

**Escalation Tool:** Patients were considered suitable for discharge from the VH pathway once all recovery domains had reached 'green' status, indicating satisfactory clinical recovery without features requiring ongoing escalation, intervention, or enhanced monitoring.

| | | |
|---|---|---|
| **Well Being & Activity** | **Risk factors** | **( √ )** |
| | Feels very unwell/rapid pulse at rest, feels cold and clammy | |
| | Has high temperature/showing signs of fever/rigors | |
| | Not mobilising/remaining in bed/extreme lethargy,unable to move | |
| | Acute breathlessness /difficulty in breathing/productive cough/chest pain | |
| | Walking occasionally but not quite resuming normal activities/ Fluctuating energy levels | |
| | Some shortness of breath and/or a productive cough | |
| | Well/active/resuming normal activities/good energy level | |
| **Pain Score** | **Risk factors** | **( √ )** |
| | Severe ongoing acute abdominal pain | |
| | Pain not controlled with regular analgesia | |
| | New increased abdominal pain since surgery | |
| | Persistent pain (anywhere) for more than 1 -2 hours | |
| | Moderate fluctuating abdominal pain | |
| | Pain not completely controlled with analgesia | |
| | Mild abdominal pain controlled with analgesia/NO pain | |
| **Nausea Vomiting & Nutrition** | **Risk factors** | **( √ )** |
| | Persistent nausea and or Vomiting | |
| | Gross abdominal distension and/or feeling bloated | |
| | Not managing/reluctant to eat & drink/no appetite with signs of dehydration | |
| | Some nausea or occasional vomiting/reflux | |
| | Managing small amounts of diet and fluids/little appetite | |
| | No nausea/vomiting and is eating and drinking | |
| **Bowel Function** | **Risk factors** | **( √ )** |
| | No Bowel motion up to 5 days/not passing flatus | |
| | Persistent diarrhoea more than 8 times a day accompanied by feeling dehydrated | |
| | PR - Pus/bleeding/offensive discharge | |
| | Bowels not yet opened up to 4 days/small amounts of flatus | |
| | Constipated stool/Some abdominal distention | |
| | Diarrhoea up to 7 times a day -otherwise well,not dehydrated | |
| | Bowels open/Passing flatus/no abdominal distention | |
| **Urinary Symptoms** | **Risk factors** | **( √ )** |
| | Extreme difficulty passing urine/retention/bladder pain | |
| | Home with a catheter? – bypassing/blocked – no support | |
| | Excessive stinging when passing urine and/or frequency | |
| | **Home catheter** - Some difficulties with the catheter and/or support services i.e. delays in treatment EG: TWOC referral | |
| | No problems passing urine | |
| | **Home catheter** - Managing catheter, TWOC referral made | |

## S2

### Virtual Hospital - Quality of life questionnaire

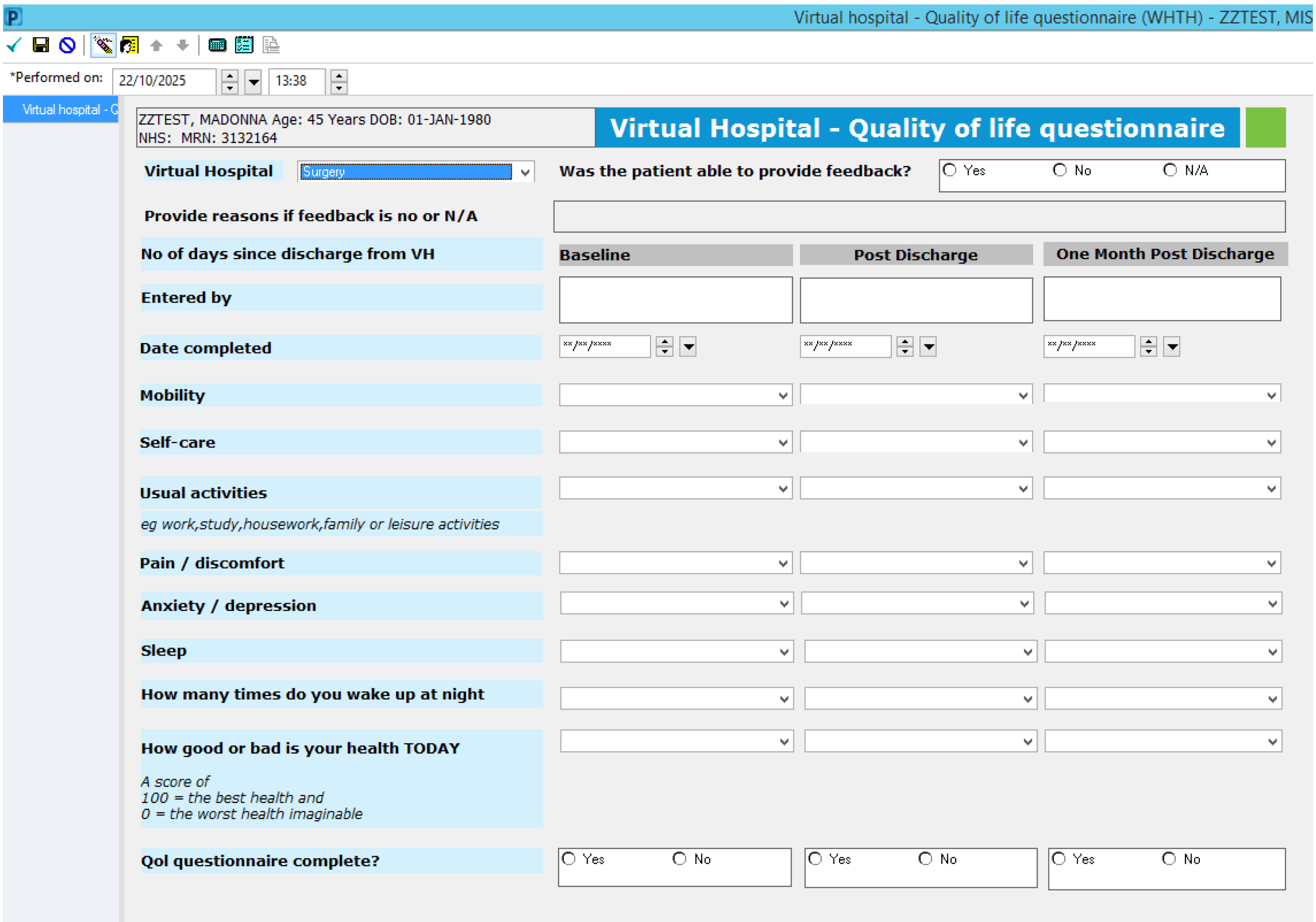

Virtual hospital - Quality of life questionnaire (WHTH) - ZZTEST, MIS

*Performed on: 22/10/2025 13:38

Virtual hospital - Q

ZZTEST, MADONNA Age: 45 Years DOB: 01-JAN-1980
NHS: MRN: 3132164

**Virtual Hospital - Quality of life questionnaire**

**Virtual Hospital** Surgery

**Was the patient able to provide feedback?** Yes No N/A

**Provide reasons if feedback is no or N/A**

| | Baseline | Post Discharge | One Month Post Discharge |
|---|---|---|---|
| **No of days since discharge from VH** | | | |
| **Entered by** | | | |
| **Date completed** | **/**/**** | **/**/**** | **/**/**** |
| **Mobility** | | | |
| **Self-care** | | | |
| **Usual activities** *eg work,study,housework,family or leisure activities* | | | |
| **Pain / discomfort** | | | |
| **Anxiety / depression** | | | |
| **Sleep** | | | |
| **How many times do you wake up at night** | | | |
| **How good or bad is your health TODAY** *A score of 100 = the best health and 0 = the worst health imaginable* | | | |
| **Qol questionnaire complete?** | Yes No | Yes No | Yes No |

### Virtual Hospital - Patient Experience Survey

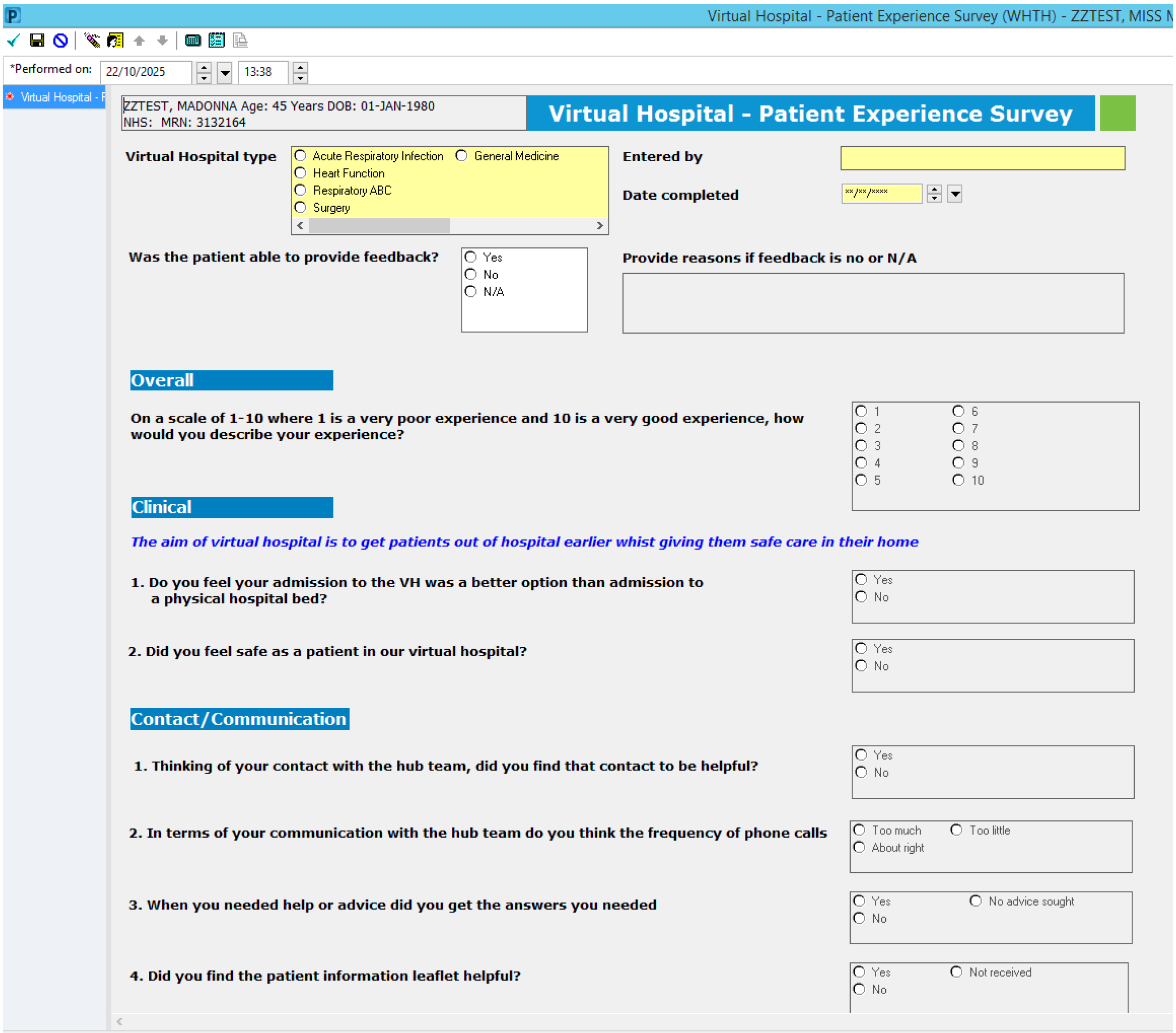

Virtual Hospital - Patient Experience Survey (WHTH) - ZZTEST, MISS M

*Performed on: 22/10/2025 13:38

Virtual Hospital - P

ZZTEST, MADONNA Age: 45 Years DOB: 01-JAN-1980
NHS: MRN: 3132164

**Virtual Hospital - Patient Experience Survey**

**Virtual Hospital type**
- ○ Acute Respiratory Infection ○ General Medicine
- ○ Heart Function
- ○ Respiratory ABC
- ○ Surgery

**Entered by**

**Date completed** **/**/****

**Was the patient able to provide feedback?**
- ○ Yes
- ○ No
- ○ N/A

**Provide reasons if feedback is no or N/A**

#### Overall

**On a scale of 1-10 where 1 is a very poor experience and 10 is a very good experience, how would you describe your experience?**
- ○ 1 ○ 6
- ○ 2 ○ 7
- ○ 3 ○ 8
- ○ 4 ○ 9
- ○ 5 ○ 10

#### Clinical

***The aim of virtual hospital is to get patients out of hospital earlier whist giving them safe care in their home***

**1. Do you feel your admission to the VH was a better option than admission to a physical hospital bed?**
- ○ Yes
- ○ No

**2. Did you feel safe as a patient in our virtual hospital?**
- ○ Yes
- ○ No

#### Contact/Communication

**1. Thinking of your contact with the hub team, did you find that contact to be helpful?**
- ○ Yes
- ○ No

**2. In terms of your communication with the hub team do you think the frequency of phone calls**
- ○ Too much ○ Too little
- ○ About right

**3. When you needed help or advice did you get the answers you needed**
- ○ Yes ○ No advice sought
- ○ No

**4. Did you find the patient information leaflet helpful?**
- ○ Yes ○ Not received
- ○ No

Virtual Hospital - Patient Experience Survey (WHTH) - ZZTEST, MIS

*Performed on: 22/10/2025 13:38

Virtual Hospital - F

**Would you like to suggest any changes?**

Segoe UI 9

**5. Did you receive a visit from one of our community teams?**

- ○ Yes ○ Unsure
- ○ No

**6. If so which .....**

- ☐ COPD ☐ District Nurse
- ☐ HF ☐ Other
- ☐ Phlebotomy
- ☐ Rapid response

**If Other community team, state**

**7. Did you find the community team helpful?**

- ○ Yes ○ Unsure
- ○ No

**8. Did you receive a call or visit from one of our partners in the voluntary section?**

- ○ Yes ○ Unsure
- ○ No

**9. If so which voluntary sector?**

**10.Did you find the voluntary sector input helpful?**

- ○ Yes ○ N/A
- ○ No

#### IT/Equipment

**1. How easy was the Massimo App to use?**

- ○ Easy ○ Difficult
- ○ Medium ○ N/A

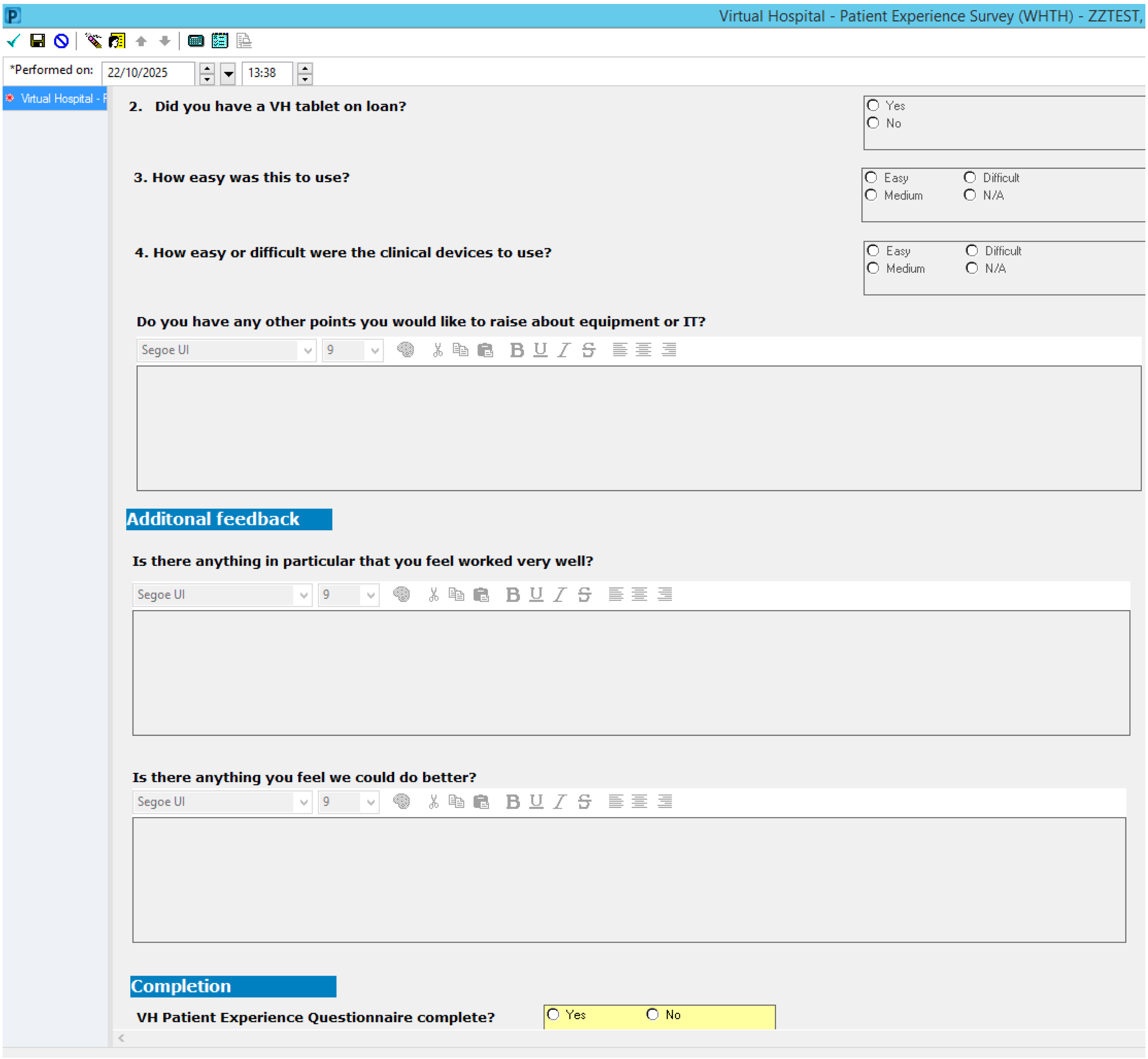

Virtual Hospital - Patient Experience Survey (WHTH) - ZZTEST,

*Performed on: 22/10/2025 13:38

Virtual Hospital - F

**2. Did you have a VH tablet on loan?**

- ○ Yes
- ○ No

**3. How easy was this to use?**

- ○ Easy
- ○ Difficult
- ○ Medium
- ○ N/A

**4. How easy or difficult were the clinical devices to use?**

- ○ Easy
- ○ Difficult
- ○ Medium
- ○ N/A

**Do you have any other points you would like to raise about equipment or IT?**

Segoe UI 9

#### Additonal feedback

**Is there anything in particular that you feel worked very well?**

Segoe UI 9

**Is there anything you feel we could do better?**

Segoe UI 9

#### Completion

**VH Patient Experience Questionnaire complete?** ○ Yes ○ No

### IQVIA™ Survey

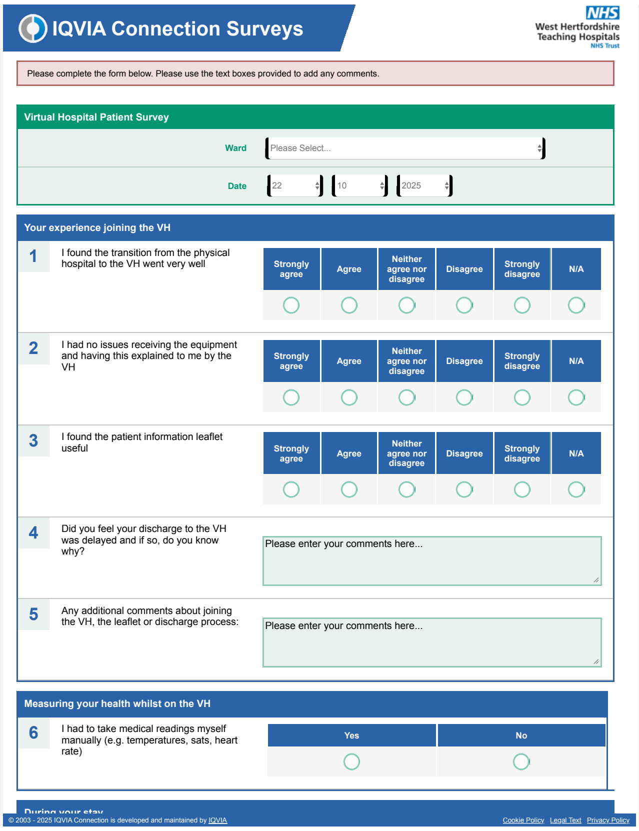

IQVIA Connection Surveys

West Hertfordshire Teaching Hospitals NHS Trust

Please complete the form below. Please use the text boxes provided to add any comments.

**Virtual Hospital Patient Survey**

Ward: Please Select...

Date: 22 / 10 / 2025

**Your experience joining the VH**

| | | Strongly agree | Agree | Neither agree nor disagree | Disagree | Strongly disagree | N/A |
|---|---|---|---|---|---|---|---|
| 1 | I found the transition from the physical hospital to the VH went very well | ○ | ○ | ○ | ○ | ○ | ○ |
| 2 | I had no issues receiving the equipment and having this explained to me by the VH | ○ | ○ | ○ | ○ | ○ | ○ |
| 3 | I found the patient information leaflet useful | ○ | ○ | ○ | ○ | ○ | ○ |

4 Did you feel your discharge to the VH was delayed and if so, do you know why?

Please enter your comments here...

5 Any additional comments about joining the VH, the leaflet or discharge process:

Please enter your comments here...

**Measuring your health whilst on the VH**

| | | Yes | No |
|---|---|---|---|
| 6 | I had to take medical readings myself manually (e.g. temperatures, sats, heart rate) | ○ | ○ |

During your stay

© 2003 - 2025 IQVIA Connection is developed and maintained by IQVIA Cookie Policy Legal Text Privacy Policy

**S3**

#### Procedure Codes

| Procedure Code | Description |
|---|---|
| G692 | Ileectomy and anastomosis of duodenum to ileum |
| G693 | Ileectomy and anastomosis of ileum to ileum |
| G694 | Ileectomy and anastomosis of ileum to colon |
| G698 | Other specified excision of ileum |
| G699 | Unspecified excision of ileum |
| G723 | Anastomosis of ileum to colon NEC |
| G724 | Anastomosis of ileum to rectum |
| G753 | Closure of ileostomy |
| H041 | Panproctocolectomy and ileostomy |
| H042 | Panproctocolectomy and anastomosis of ileum to anus and creation of pouch HFQ |
| H053 | Total colectomy and ileostomy NEC |
| H062 | Extended right hemicolectomy and anastomosis of ileum to colon |
| H063 | Extended right hemicolectomy and anastomosis NEC |
| H064 | Extended right hemicolectomy and ileostomy HFQ |
| H071 | Right hemicolectomy and end to end anastomosis of ileum to colon |
| H072 | Right hemicolectomy and side to side anastomosis of ileum to transverse colon |
| H073 | Right hemicolectomy and anastomosis NEC |
| H074 | Right hemicolectomy and ileostomy HFQ |
| H075 | Right hemicolectomy and end to side anastomosis |
| H078 | Other specified other excision of right hemicolon |
| H079 | Unspecified other excision of right hemicolon |
| H085 | Transverse colectomy and exteriorisation of bowel NEC |
| H091 | Left hemicolectomy and end to end anastomosis of colon to rectum |
| H092 | Left hemicolectomy and end to end anastomosis of colon to colon |
| H093 | Left hemicolectomy and anastomosis NEC |
| H095 | Left hemicolectomy and exteriorisation of bowel NEC |
| H096 | Left hemicolectomy and end to side anastomosis |
| H098 | Other specified excision of left hemicolon |
| H101 | Sigmoid colectomy and end to end anastomosis of ileum to rectum |
| H102 | Sigmoid colectomy and anastomosis of colon to rectum |
| H103 | Sigmoid colectomy and anastomosis NEC |
| H104 | Sigmoid colectomy and ileostomy HFQ |
| H105 | Sigmoid colectomy and exteriorisation of bowel NEC |
| H112 | Colectomy and side to side anastomosis of ileum to colon NEC |
| H114 | Colectomy and ileostomy NEC |
| H154 | Closure of colostomy |
| H295 | Subtotal excision of colon and anastomosis of colon to ileum |
| H298 | Other specified subtotal excision of colon |
| H299 | Unspecified subtotal excision of colon |
| H331 | Abdominoperineal excision of rectum and end colostomy |

| | |
|---|---|
| H332 | Proctectomy and anastomosis of colon to anus |
| H333 | Anterior resection of rectum and anastomosis of colon to rectum using staples |
| H334 | Anterior resection of rectum and anastomosis NEC |
| H335 | Rectosigmoidectomy and closure of rectal stump and exteriorisation of bowel |
| H336 | Anterior resection of rectum and exteriorisation of bowel |
| H337 | Perineal resection of rectum HFQ |
| H338 | Other specified excision of rectum |
| H339 | Unspecified excision of rectum |
| H341 | Open excision of lesion of rectum |
| H411 | Rectosigmoidectomy and peranal anastomosis |

#### Supplementary Results

### S4

**Table S4** summarises the ethnic composition between VH and Non-VH cohorts. The majority of patients were White British (VH = 56; Non-VH=199). There was no statistically significant difference in the proportion of White versus minority ethnic patients between groups (p=0.396).

| Ethnicity | VH | Non-VH |
|---|---|---|
| White – British | 56 (69) | 199 (79) |
| White – Any Other White Background | 7 (9) | 13 (5) |
| White – Irish | 3 (4) | 6 (2) |
| Asian – Indian | 1 (1) | 5 (2) |
| Asian – Bangladeshi | 0 (0) | 3 (1) |
| Asian – Other Asian Background | 1 (1) | 3 (1) |
| Pakistani | 0 (0) | 1 (0) |
| Black – Caribbean | 1 (1) | 3 (1) |
| Black – Other Black Background | 0 (0) | 3 (1) |
| Mixed – Any Other Mixed Background | 0 (0) | 2 (1) |
| Chinese | 0 (0) | 1 (0) |
| Other – Any Other Ethnic Group | 0 (0) | 3 (1) |
| Not Stated | 12 (15) | 11 (4) |
